# Reproducibility of COVID-era infectious disease models

**DOI:** 10.1101/2023.10.11.23296911

**Authors:** Alec S. Henderson, Roslyn I. Hickson, Morgan Furlong, Emma S. McBryde, Michael T. Meehan

**Affiliations:** Australian Institute of Tropical Health and Medicine, James Cook University, Townsville, Australia; College of Public Health, Medical and Veterinary Sciences, James Cook University, Townsville, Australia; Commonwealth Scientific Industrial Research Organisation (CSIRO), Townsville, Australia

**Keywords:** Infectious disease modelling, COVID-19, Open science, Reproducibility

## Abstract

Infectious disease modelling has been prominent throughout the COVID-19 pandemic, helping to understand the virus’ transmission dynamics and inform response policies. Given their potential importance and translational impact, we evaluated the computational reproducibility of infectious disease modelling articles from the COVID era. We found that only four out of 100 randomly sampled studies released between January 2020 and August 2022 could be computationally reproduced using the resources provided (e.g., code, data, instructions). For the 100 most highly cited articles from the same period we found that only 11 were reproducible. Reflecting on our experience, we discuss common issues affecting computational reproducibility and how these might be addressed.

## Introduction

Mathematical models are useful tools for analysing infectious disease outbreaks. They have been used to elucidate mechanisms of spread [1], facilitate the design and evaluation of different control strategies [2] and guide public health decision-making [3]. Never has this been more evident than with the COVID-19 pandemic, where modelling has helped to characterise the biological and epidemiological properties of the aetiological agent (SARS-CoV-2) and its associated disease (COVID-19) [4, 5, 6]. Modelling has also been used to forecast epidemic trajectories under various scenarios [7, 8], supporting decisions to implement or revoke particular interventions – e.g., travel restrictions, social distancing.

Whilst public health policy is typically informed by numerous factors (e.g., social, economic), modelling is increasingly recognised as a valuable decision support tool. Given this responsibility, policy-relevant studies should be clearly described, transparent and reproducible [9]. This allows external reviewers to readily critique: the quality and relevance of input data; the design and implementation of model structures; and the validity and robustness of modelling assumptions, among others. Within this, the central role of numerical simulation and computation in infectious disease modelling makes the provision of working code underpinning computational results necessary.

Recently, Zavalis et al. [10] evaluated the transparency of published articles in infectious disease modelling by recording the number of papers that had publicly released accompanying code. They found that few authors (20%) share their code, with Collins et al. [11] obtaining similar results for COVID-19 preprints (21-33%). These release rates align with other computational domains such as machine learning (33%) [12] and physics (6%) [13].

Despite this, transparency is still only one element of reproducible research. Providing source code that either does not work or produces inconsistent results may counter reproducibility. Nevertheless, reproducibility studies scarcely probe beyond presence/absence evaluations and check that the code that is made available meets its desired purpose. So, whilst computational reproducibility is widely accepted as a systemic problem plaguing quantitative research [14], the extent to which infectious disease modelling articles fail in this regard and the specific reasons why remain unquantified.

In this work, we assess the reproducibility of contemporary infectious disease modelling studies by determining the extent to which quantitative results can be reproduced using the computational materials provided (e.g., code, data). Further, to help understand the barriers to computational reproducibility and possible resolutions, we investigate whether practices such as journal mandates, the provision of instructions, or notebook-style formatting are associated with increased reproducibility.

## Methods

### Literature search

We chose Google Scholar as the search repository so as not to discriminate against preprints (many of which are highly cited and have been impactful throughout the pandemic). To limit our investigation to articles in the COVID era, we conducted multiple searches of articles released from January 2020 to August 2022 using the terms: ‘SARS-CoV-2 model*’; ‘infectious disease model* covid’, ‘infectious disease model*’; ‘coronavirus model*’; ‘covid model*’; ‘Infectious disease modeling’; ‘Infectious disease modelling’ and ‘Infectious disease simulations’. Due to Google Scholar restricting each search to return at most 1,000 results, we used many similar search terms to obtain a more complete data set. We repeated the searches on two different machines using private browsers to minimise the effects of randomness and user bias. We also recorded the citation count of each article as reported by Google Scholar (as of August 2022).

### Processing the search results

Combining each of the 1,000 results returned from these separate searches, we analysed two subsets: the 100 most cited and a random sample of 100 papers (acknowledging these subsets may intersect). To generate these subsets, we screened for papers that contained an original simulation or computational component relating to infectious disease transmission dynamics and for which *any* results would be reasonably assumed to be made using computational software (papers with minor computations were still included if the paper contained new software-generated results). Conversely, we excluded all: reviews; commentaries; non-original research articles; theses; and papers written in a language other than English. The list of included/excluded papers was independently checked by two authors, with discrepancies settled by joint consultation with all authors. A diagram giving a high-level representation of this process is presented in Figure 1.

**Figure 1:**
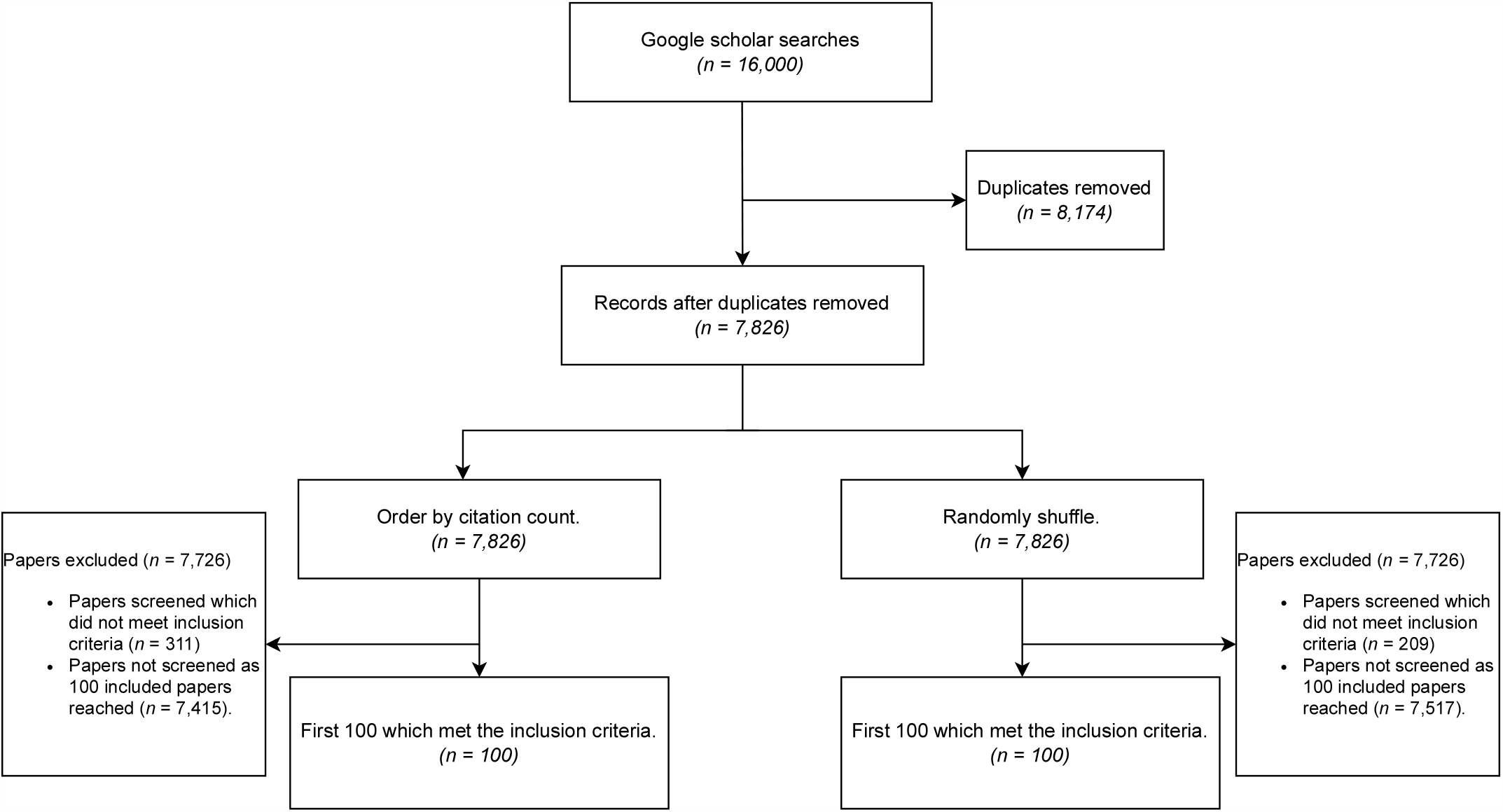
A diagram for papers to be included in our analysis.

### Evaluating the papers

The evaluation process for each paper can be divided into three steps. First, we assessed the transparency and inclusion of code-independent information in the papers. This involved documenting: the presence of data; the availability of code (e.g. through a functional link to the *source* code); the journal where the article was published; whether said journal mandated statements on data or code availability (in this context, a mandate refers to a requirement for a statement or section in the document which is not described as optional); and any institutional affiliations of the authors.

Second, where a link to the code was provided, we noted the details of the code and storage location. We recorded: the programming language used; whether the programming environment was open source; and whether the code was presented in a notebook format. For interest we also noted: the licensing details for the code; whether the repository was self-contained; and whether the version of the code for the paper was archived (will not change).

Finally, we evaluated the reproducibility of the results, recording: whether all/some of the figures, tables or numerical results appearing in the main paper were reproduced using available resources; and whether the code was cross-platform (would run on both Windows and Linux).

For *each* paper, two *new* virtual machines were created and used to ensure a fair comparison and that the testing environment was as pristine as possible: one virtual machine running the Windows 11 development environment and the other Ubuntu version 22.04. We used virtual box to run the virtual machines as it is a cross-platform virtualisation software. For each paper, we followed all given instructions on setting up the environment. If no information was given, we used the latest version of the software (unless it was clear to use a different version) required to run the code. The order in which we ran scripts was determined by instructions, the naming of files or our basic understanding of the code given. If, during the running of the code, an error occurred, we reviewed the error message given to determine if it was simply caused by misplaced files, the naming of paths, or some other very minor issue (e.g., failure to load a required package or clearing pre-generated results); in which case we resolved the issue and reran the code.

We counted code as outputting all results of the paper if all the graphs and tables/numbers presented in the main article were reproduced. We counted code as outputting some results if it produced at least one graph or number presented in the article but not all. If the code was non-deterministic or used different data from the original study, we counted it as reproducing all or some of the results if this was deemed to cause the differences. We assessed graphs based on their numeric content, ignoring slight differences in presentation.

We placed no limit on the amount of time for reproducing the results of studies (computational time ranged from a few seconds to weeks). When resources such as proprietary software/data were not able to be sourced (that is, were unavailable to the authors through any of their institutional subscriptions or free trials) and were required, the article was deemed to not be reproducible (noting that this may not mean the entire study is not reproducible).

### Statistics

Logistic regression was used to assess the effect of data mandates, code mandates, and log transformed citation count on the proportion of articles that release code or data, referred to hereafter as the release rate. A multivariate analysis was run to test for significance, including only the input variables with a univariate *p <* 0.1 as discussed in [15]. The results for the univariate and multivariate analyses are presented in Tables 3, 4, and 5.

**Table 1:** Characteristics of infectious disease modelling articles.^*†*^Articles that have been published in peer-reviewed journals.^*\**^Inter-quartile range.# Note that the random and top-cited sets overlap, containing a total of 193 papers; see the available data for more details.

|  | Random | Top cited |
| --- | --- | --- |
| **Papers** | 100 | 100 |
| Peer reviewed<sup>†</sup>, $n$ | 91 | 95 |
| Citations, median (IQR*) | 21 (5 - 47) | 653 (480 - 1072) |
| **Journal Mandate** |  |  |
| Data availability, $n$ | 43 | 71 |
| Code availability, $n$ | 11 | 25 |
| Data and Code availability, $n$ | 9 | 25 |
| **Disease** |  |  |
| COVID, $n$ | 76 | 100 |
| Other, $n$ | 24 | 0 |
| **Link provided** |  |  |
| Data, $n$ | 67 | 80 |
| Code, $n$ | 19 | 48 |
| Code and Data, $n$ | 19 | 48 |

**Table 2:**
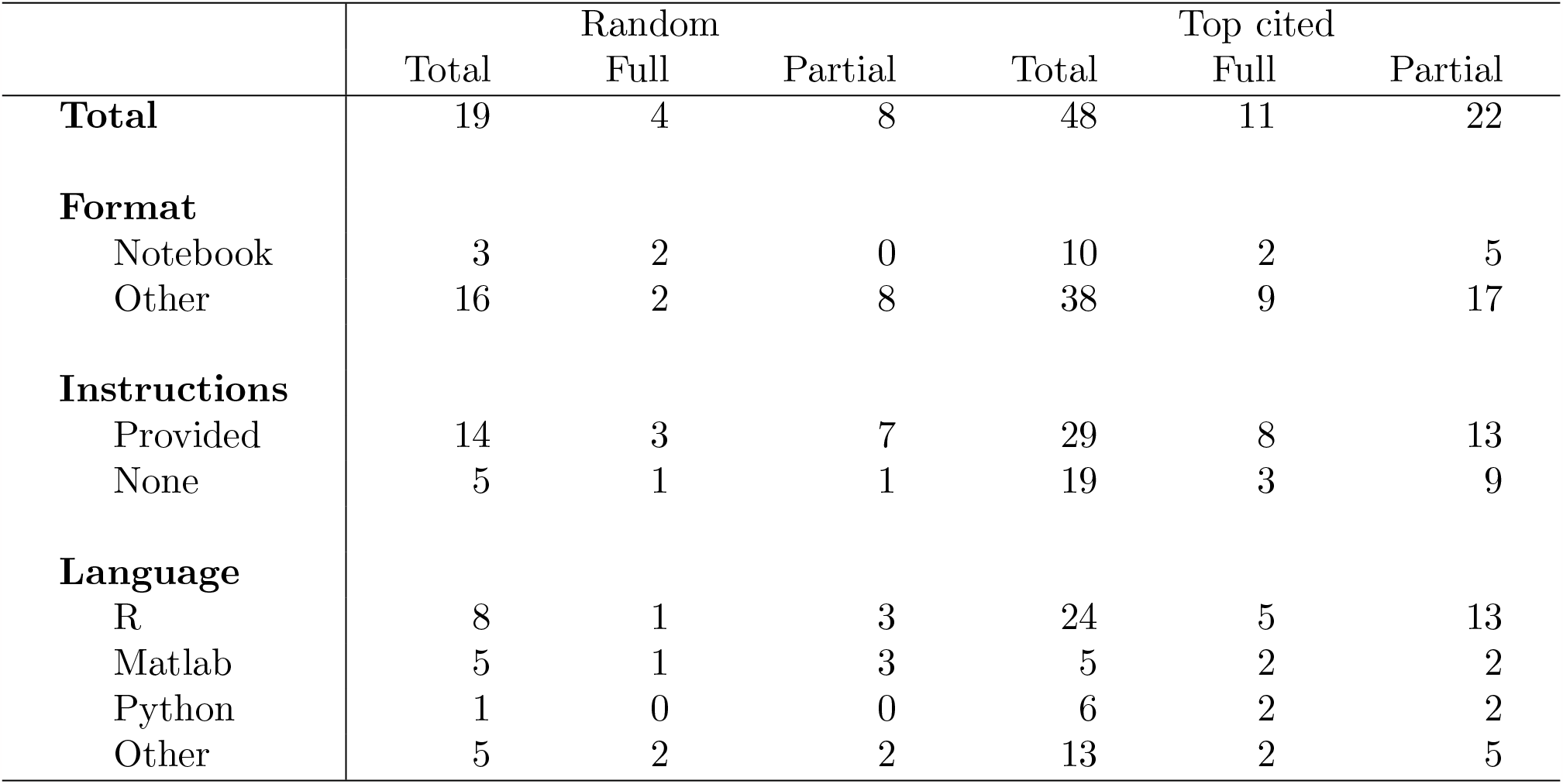
Reproducibility of papers for which code was provided.

**Table 3:** Effect size results from univariate logistic regression applied to transparency outcomes. For each binary variable, the absence of the given covariate is taken as the reference level (e.g., no data mandate). OR: Odds ratio. CI: Confidence interval. ^*†*^ The citation count was log transformed.

|  | Random |  |  |  | Top cited |  |  |  |
| --- | --- | --- | --- | --- | --- | --- | --- | --- |
| | OR (95% CI) | LR$\chi^2$ | df | *p* | OR (95% CI) | LR$\chi^2$ | df | *p* |
| **Data release** |  |  |  |  |  |  |  |  |
| Data mandate | 1.51 (0.65 - 3.62) | 0.89 | 1 | 0.35 | 1.42 (0.48 - 4.00) | 0.43 | 1 | 0.51 |
| Code mandate | 0.55 (0.15 - 2.05) | 0.83 | 1 | 0.36 | 0.40 (0.14 - 1.18) | 2.79 | 1 | 0.10 |
| Citations | 1.40 (1.07 - 1.88) | 6.14 | 1 | 0.01 | 6.84 (1.98 - 35.9) | 11.2 | 1 | < 0.01 |
| **Code release** |  |  |  |  |  |  |  |  |
| Data mandate | 5.02 (1.73 - 16.8) | 9.09 | 1 | < 0.01 | 2.71 (1.11 - 7.03) | 4.81 | 1 | 0.03 |
| Code mandate | 2.82 (0.67 - 10.6) | 2.09 | 1 | 0.15 | 1.91 (0.77 - 4.92) | 1.93 | 1 | 0.17 |
| Citations | 1.30 (0.96 - 1.79) | 2.92 | 1 | 0.09 | 2.31 (1.22 - 4.74) | 6.69 | 1 | 0.01 |

**Table 4:** Effect size results from multivariate logistic regression applied to transparency outcomes for including covariates with *p* values *<* 0.10 from the univariate analysis. For each binary variable, the absence of the given covariate is taken as the reference level (e.g., no data mandate). OR: Odds ratio. CI: Confidence interval. ^*†*^ The citation count was log transformed.

|  | Random |  |  |  |
| --- | --- | --- | --- | --- |
| | OR (95% CI) | LR$\chi^2$ | df | *p* |
| **Data release** |  |  |  |  |
| Citations | 1.40 (1.07 - 1.88) | 6.14 | 1 | 0.01 |
| **Code release** |  |  |  |  |
| Data mandate | 4.39 (1.43 - 15.4) | 6.74 | 1 | 0.01 |
| Citations | 1.13 (0.82 - 1.58) | 0.57 | 1 | 0.44 |

**Table 5:**
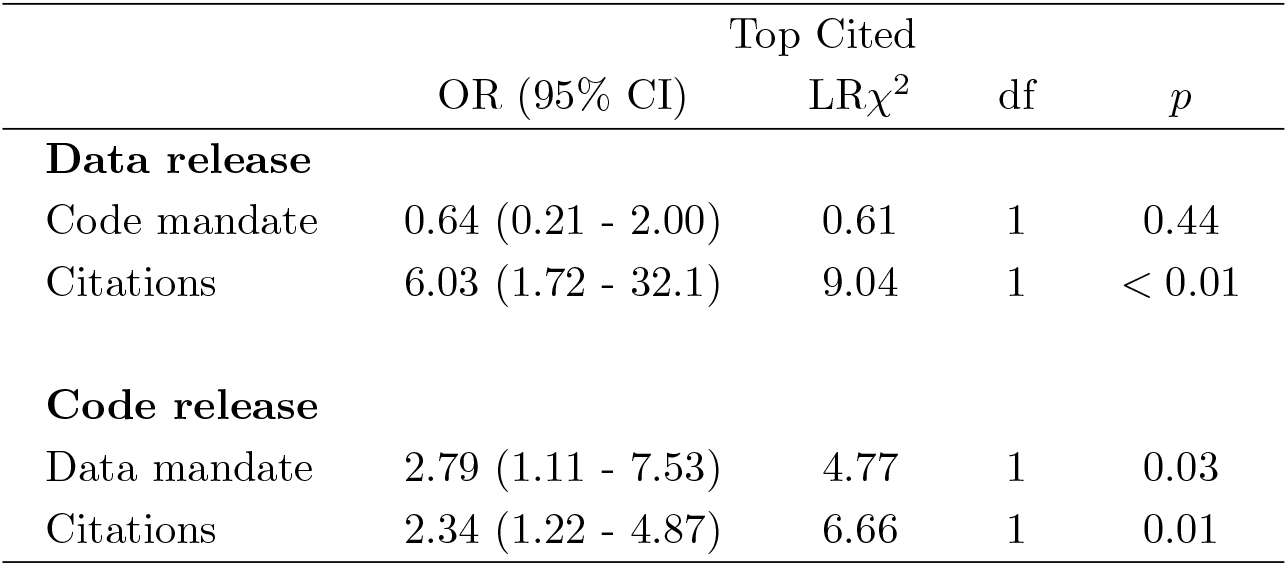
Effect size results from multivariate logistic regression applied to transparency outcomes for including covariates with *p* values *<* 0.10 from the univariate analysis. For each binary variable, the absence of the given covariate is taken as the reference level (e.g., no data mandate). OR: Odds ratio. CI: Confidence interval. ^*†*^ The citation count was log transformed.

|  | Top Cited |  |  |  |
| --- | --- | --- | --- | --- |
| | OR (95% CI) | LR$\chi^2$ | df | *p* |
| **Data release** |  |  |  |  |
| Code mandate | 0.64 (0.21 - 2.00) | 0.61 | 1 | 0.44 |
| Citations | 6.03 (1.72 - 32.1) | 9.04 | 1 | < 0.01 |
| **Code release** |  |  |  |  |
| Data mandate | 2.79 (1.11 - 7.53) | 4.77 | 1 | 0.03 |
| Citations | 2.34 (1.22 - 4.87) | 6.66 | 1 | 0.01 |

Similarly, univariate ordinal regression was used to assess the effect of instructions, code format (notebook or other), the programming language used, and number of log transformed citations on the level of reproducibility (see Table 6). In the event that the assumptions for ordinal regression were not met (i.e., proportional odds did not hold), we used multinomial regression.

**Table 6:** Effect size results from univariate ordinal regression applied to reproducibility outcomes. For each binary variable, the absence of the given covariate is taken as the reference level (e.g., no instructions). OR: Odds ratio. CI: Confidence interval.^*#*^ Results from multinomial regression as the proportional odds assumption was violated, we present the odds ratio for complete vs no reproducibility. ^*†*^ The citation count was log transformed. * For the random group, the Python language (*n* = 1) was grouped with Other languages.

|  | Random |  |  |  | Top cited |  |  |  |
| --- | --- | --- | --- | --- | --- | --- | --- | --- |
| | OR (95% CI) | LR$\chi^2$ | df | *p* | OR (95% CI) | LR$\chi^2$ | df | *p* |
| Instructions | 2.73 (0.36 - 26.4) | 0.94 | 1 | 0.33 | 1.70 (0.58 - 5.16) | 0.93 | 1 | 0.34 |
| Notebook | 6.00 (0.34 - 107)<sup>#</sup> | 5.29<sup>#</sup> | 2<sup>#</sup> | 0.07<sup>#</sup> | 0.82 (0.22 - 3.00) | 0.09 | 1 | 0.77 |
| Citations<sup>†</sup> | 1.06 (0.70 - 1.63) | 0.09 | 1 | 0.77 | 1.19 (0.60 - 2.40) | 0.25 | 1 | 0.62 |
| Language | — | 1.22 | 2 | 0.54 | — | 2.26 | 3 | 0.52 |
| *Matlab* | 1 (reference) | - | - | - | 1 (reference) | - | - | - |
| *R* | 0.38 (0.11 - 9.40) | - | - | - | 0.52 (0.08 - 3.33) | - | - | - |
| *Python* | * | - | - | - | 0.58 (0.05 - 5.91) | - | - | - |
| *Other* | 1.00 (0.11 - 9.40) | - | - | - | 0.25 (0.03 - 1.86) | - | - | - |

For the effect size of each variable, we report the maximum likelihood estimate of the odds ratio (OR) alongside its 95% confidence interval. Further, we conducted likelihood ratio tests to evaluate the statistical significance of each covariate considered.

The code and data required to reproduce our analysis are available at: https://github.com/AITHM/Reproducing-the-Infectious-Disease-Models-of-the-Covid-Era.

## Results

Our literature search yielded 16,000 results, of which 7,826 were unique. To generate two sets of 100 eligible articles (totalling 193 unique articles), we screened 309 and 411 papers for the random and top cited groups, respectively. The characteristics of these articles are summarised in Table 1.

For interest, we note that all of the 100 most highly cited articles were COVID-related (compared with 76 of the random set) and had a median (IQR) citation count of 653 (480 - 1,072).

Data links were available for 67 of the 100 randomly sampled articles and 80 of the top cited. In contrast, code links were provided 19 and 48 times for each group, respectively. Surprisingly, only 18 of the 34 articles published in journals that mandate code availability provided a working link to their source code. We found no significant association between data release rates and data mandates (*p* = 0.35; top cited: *p* = 0.51) and code release rates and code mandates (random: *p* = 0.15; top cited: *p* = 0.17). However, we did observe significant associations for both groups between code release rates and data mandates (random: OR 4.39, 95% CI 1.43 to 15.4, *p* = 0.01, top cited: OR 2.79, 95% CI 1.11 to 7.53, *p* = 0.03) and log transformed citation count and data release (random: OR 1.40, 95% CI 1.07 to 1.88, *p* = 0.01; top cited: OR 6.03, 95% CI 1.72 - 32.1, *p <* 0.01). Finally we also found a significant association between log transformed citation count and code release in the top-cited group (OR 2.34, 95% CI 1.22 to 4.87, *p* = 0.01). See Tables 3, 4, and 5 for more details.

For those articles that did release their code, we found R to be the most popular programming language used (featuring 50% of the time for the top cited and 42% for the random set), followed by Matlab.

Running the code provided, we found low levels of reproducibility for both groups, as summarised in Table 2. In particular, *only 4 of the 19* randomly sampled articles were fully reproducible, whilst 8 were partially reproducible (i.e., we could reproduce at least one result, but not all results). Alternatively, for the most highly cited group we found that 11 articles were fully reproducible, whilst a further 22 were partially reproducible. One paper was counted as not being reproducible due to being unable to obtain proprietary data. We found that where code was provided, the likelihood of reproducing results was not associated with: the provision of instructions (random: *p* = 0.33; top cited: *p* = 0.34); the use of notebooks (random: *p* = 0.07; top: *p* = 0.77); the log transformed citation count (random: *p* = 0.77; top: *p* = 0.62); or the programming language used (random: *p* = 0.54; top: *p* = 0.52), see Table 6 for more details.

With the most popular languages used being cross-platform (although some extensions may not be), we found that the vast majority of code provided ran on both Windows and Linux (at least up to the same point if an error occurred).

## Discussion

In this study we verified previous code transparency results [10, 11], finding that less than 20% of randomly selected infectious disease modelling studies provide working links to code. Notably, the top-cited set of studies released code far more frequently (48%); however this is may be a function of the journals in which these articles were published and the mandates which they impose. In any case, code release rates still considerably lag data release rates (67% random; 80% top-cited).

In our main analysis, we extended previous investigations by testing the reproducibility of code that was made available. Notably, only four out of the 19 randomly sampled studies that provided code were found to be *completely* computationally reproducible, translating to an overall reproducibility rate of 4%. Whilst the code from a further eight studies could reproduce at least one correct numerical value or figure, the results are still disappointing. For the top-cited articles, the reproducibility rates of released code are similar (11 out of 48 were completely reproducible, and 22 out of 48 were partially reproducible); however, the higher rates of code release in this group yielded much greater reproducibility rates overall (11% complete; 22% partial).

For context, one of the few similar computational reproducibility studies, which was conducted in computational physics, found that none of the 55 articles considered were reproducible [13]. However, in contrast to our study, the authors of [13] imposed strict time limits on computation and focused on papers from a single journal. Similarly we note a source of bias in our own study is for the top cited (random) articles, the three most prominent affiliations are co-authors on 23% (15%) of the papers, whereas the top five co-author 35% (22%).

The low rates of code release may be a consequence of several factors which have been discussed previously [16]: authors perceiving their code as inelegant or unwieldy; concerns over proprietary rights; or potential maintenance obligations associated with publicly available code. However, in the context of the COVID-19 pandemic, time constraints were presumably a significant limiting factor. Still, data release rates from this period remain much higher, indicating that the priority that is placed on data availability has not yet been translated to code — something that is mirrored by the number of journals that impose data and/or code release mandates.

For papers that did release code, there were many reasons why results were not reproduced. Often, the code was incomplete, either: generating only subsets of published outputs (e.g., failing to produce any figures); or omitting private dependencies (e.g., custom scripts). Some studies only provided code examples, designed to demonstrate the methodology, not to functionally reproduce results. Whilst very rare, some papers’ code executed successfully, but the generated outputs diverged from published results.

Even when articles were found to be at least partially reproducible, some intervention was typically needed. Required packages and dependencies were frequently not loaded, or file paths were referenced incorrectly (e.g., using absolute rather than relative referencing). However, these issues were often easily corrected, and the articles were subsequently assessed without penalty.

When multiple scripts were provided, unclear or limited instructions made it difficult to determine the appropriate computation order – a problem that was amplified for larger repositories with intricate dependencies. When unsure, we systematically permuted the order of computation (to the extent that this was feasible); however, the success of this strategy varied markedly with repository size.

Our results show that the programming language used, formatting structure (e.g., notebooks), and provision of instructions were not statistically significantly associated with increased rates of computational reproducibility. However, given the low rates of transparency and computational reproducibility overall, it is likely that more data is required before conclusions can be reached.

Despite these results, we found that codebases with logical formatting and detailed documentation were more straightforward to evaluate. Code written in a notebook format was typically self-documenting and mimicked the presentation of the accompanying article. Even when these studies were found not to be computationally reproducible, they are arguably more reproducible in the broader sense, in that the methodology and implementation were considerably more interpretable.

We recognise that many different criteria have been proposed for evaluating and improving reproducibility (see, e.g., [13, 17, 18]). Whilst the criteria we use follows the same intent, we have tried to minimise subjectivity and favour demonstrable reproducibility over adherence to rigid reporting standards. For example, in our view, a program with clearly named files and extensive self-documentation should not be penalised for omitting less critical details such as the operating system version number on which it was run. We emphasise that many of the computational reproducibility issues we recorded (e.g., incompleteness, run-time errors) were identified because an independent party ran the code in a fresh environment. Therefore, we strongly recommend that authors consult colleagues or external individuals to test their code, checking that it runs without error, and successfully reproduces *all* results. We also suggest that authors create a clean environment for the code themselves, e.g., using virtualisation software such as containerisation, virtual machines, or a code-running service like Google Colab or GitHub actions. The public release of the environment will also prolong the life of the software (e.g., after some required packages are no longer available) and make it easier for others to use, adapt and extend. Alternatively, editors may request that reviewers (i) check for the presence of code when appropriate (enforcing existing code release mandates); and (ii) ensure that it successfully reproduces all computational results.

Whilst our results clearly demonstrate that infectious disease modelling lacks computational reproducibility, we note that this is only one element of reproducible research. Transparency, replicability and robustness are all important metrics to evaluate a particular study’s validity. Nevertheless, computational reproducibility remains an important first step towards verification; and it has been argued that not releasing reproducible code is tantamount to stating a mathematical theorem without proof [19].

## Conclusion

Infectious disease modelling, as with other computational fields, has low computational reproducibility. We have found that journals mandating data release are significantly associated with code release. Hence, an option moving forward is more journal mandates in data and code release. However, given that many papers were published in code-mandated journals and still did not provide complete, working code, there needs to be a change in expectations from editors, reviewers, and authors. The infectious disease modelling community should make a concerted effort to release complete working code alongside appropriate data and instructions. Not doing so conceals assumptions and prevents thorough peer review. Finally, publishing code leaves a legacy and benefits all by facilitating reuse and improvement, enabling more rapid scientific progress.

## Data Availability

Anonymised data and code for reproducing the claims and table of the paper can be found at:
https://github.com/AITHM/Reproducing-the-Infectious-Disease-Models-of-the-Covid-Era

## Data availability

Anonymised data and code for reproducing the claims and table of the paper can be found at: https://github.com/AITHM/Reproducing-the-Infectious-Disease-Models-of-the-Covid-Era

## Acknowledgements

The authors would like to thank Dr. Joseph Moxon and Prof. Rhondda Jones for helpful discussions and suggestions. M.M. is supported by an Australian Research Council Discovery Early Career Researcher Award, DE210101344.

## References

[1] A. Kucharski, C. L. Althaus, The role of superspreading in Middle East respiratory syndrome coronavirus (MERS-CoV) transmission, Eurosurveillance 20 (25) (2015) 21167. doi:10.2807/1560-7917.ES2015.20.25.21167.

[2] M. Lipsitch, T. Cohen, B. Cooper, J. M. Robins, S. Ma, L. James, G. Gopalakrishna, S. K. Chew, C. C. Tan, M. H. Samore, et al., Transmission dynamics and control of severe acute respiratory syndrome, science 300 (5627) (2003) 1966–1970.

[3] D. L. Smith, K. E. Battle, S. I. Hay, C. M. Barker, T. W. Scott, F. E. McKenzie, Ross, macdonald, and a theory for the dynamics and control of mosquito-transmitted pathogens, PLoS pathogens 8 (4) (2012) e1002588.

[4] S. A. Lauer, K. H. Grantz, Q. Bi, F. K. Jones, Q. Zheng, H. R. Meredith, A. S. Azman, N. G. Reich, J. Lessler, The incubation period of coronavirus disease 2019 (covid-19) from publicly reported confirmed cases: estimation and application, Annals of internal medicine 172 (9) (2020) 577–582.

[5] R. Li, S. Pei, B. Chen, Y. Song, T. Zhang, W. Yang, J. Shaman, Substantial undocumented infection facilitates the rapid dissemination of novel coronavirus (sars-cov-2), Science 368 (6490) (2020) 489–493.

[6] R. Verity, L. C. Okell, I. Dorigatti, P. Winskill, C. Whittaker, N. Imai, G. Cuomo-Dannenburg, H. Thompson, P. G. Walker, H. Fu, et al., Estimates of the severity of coronavirus disease 2019: a model-based analysis, The Lancet infectious diseases 20 (6) (2020) 669–677.

[7] R. M. Anderson, H. Heesterbeek, D. Klinkenberg, T. D. Hollingsworth, How will country-based mitigation measures influence the course of the covid-19 epidemic?, The lancet 395 (10228) (2020) 931–934.

[8] N. Ferguson, D. Laydon, G. Nedjati Gilani, N. Imai, K. Ainslie, M. Baguelin, S. Bhatia, A. Boonyasiri, Z. Cucunuba Perez, G. Cuomo-Dannenburg, et al., Report 9: Impact of non-pharmaceutical interventions (NPIs) to reduce COVID19 mortality and healthcare demand (2020).

[9] R. Bromme, N. G. Mede, E. Thomm, B. Kremer, R. Ziegler, An anchor in troubled times: Trust in science before and within the covid-19 pandemic, PLoS One 17 (2) (2022) e0262823.

[10] E. A. Zavalis, J. P. Ioannidis, A meta-epidemiological assessment of transparency indicators of infectious disease models, Plos one 17 (10) (2022) e0275380.

[11] A. Collins, R. Alexander, Reproducibility of COVID-19 pre-prints, Scientometrics 127 (8) (2022) 4655–4673.

[12] M. B. McDermott, S. Wang, N. Marinsek, R. Ranganath, L. Foschini, M. Ghassemi, Reproducibility in machine learning for health research: Still a ways to go, Science Translational Medicine 13 (586) (2021).

[13] V. Stodden, M. S. Krafczyk, A. Bhaskar, Enabling the verification of computational results: An empirical evaluation of computational reproducibility, in: Proceedings of the First International Workshop on Practical Reproducible Evaluation of Computer Systems, 2018, pp. 1–5.

[14] R. Reinecke, T. Trautmann, T. Wagener, K. Schüler, The critical need to foster computational reproducibility, Environmental Research Letters 17 (4) (2022) 041005.

[15] P. Ranganathan, C. Pramesh, R. Aggarwal, Common pitfalls in statistical analysis: logistic regression, Perspectives in clinical research 8 (3) (2017) 148.

[16] R. J. LeVeque, Top ten reasons to not share your code (and why you should anyway), Siam News 46 (3) (2013) 15.

[17] S. Pollett, M. A. Johansson, N. G. Reich, D. Brett-Major, S. Y. Del Valle, S. Venkatramanan, R. Lowe, T. Porco, I. M. Berry, A. Deshpande, et al., Recommended reporting items for epidemic forecasting and prediction research: The epiforge 2020 guidelines, PLoS medicine 18 (10) (2021) e1003793.

[18] D. Pokutnaya, B. Childers, A. E. Arcury-Quandt, H. Hochheiser, W. G. Van Panhuis, An implementation framework to improve the transparency and reproducibility of computational models of infectious diseases, PLOS Computational Biology 19 (3) (2023) e1010856.

[19] J. B. Buckheit, D. L. Donoho, Wavelab and reproducible research, Springer, 1995.

